# Frequent testing regimen based on salivary samples for an effective COVID-19 containment strategy

**DOI:** 10.1101/2020.10.13.20210013

**Authors:** Mario Plebani, Ada Aita, Anna Maria Cattelan, Francesco Bonfante, Andrea Padoan, Carlo Giaquinto, Daniela Basso

## Abstract

Rapid and accurate diagnostic tests are essential for controlling the ongoing COVID-19 pandemic. Although the current gold standard involves testing of nasopharyngeal swabs specimens by nucleic acid amplification test, such as real-time reverse-transcription polymerase chain reaction (rRT-PCR) to detect the severe acute respiratory syndrome coronavirus 2 (SARS-CoV-2), it presents several limitations that ultimately may translate into a bottleneck in the surveillance regimen. New strategies based on frequent testing using less invasive specimens are urgently needed for containment of the infection. Rapid antigen assay using saliva as a reliable alternative to nasopharyngeal swabs should be proposed as a valuable part of the overall testing strategy.

## Introduction

Coronavirus disease 2019 (COVID-19), caused by the severe acute respiratory syndrome coronavirus 2 (SARS-CoV-2), is a serious and potentially deadly disease that spread very quickly across the world and was declared a pandemic by the World Health Organization (WHO) in March 2020. During the first wave of the pandemic, all health care systems have been coped with major challenges including the need for a more rapid and accurate diagnosis of the infection. In order to trace the disease and to implement strategies aimed at breaking the chain of transmission, WHO recommended extensive testing for SARS-CoV-2, since the contribution of presymptomatic and asymptomatic individuals to overall SARS-CoV-2 transmission has been demonstrated (1-3). This is particularly important considering the current “second” wave of the epidemic which is mostly sustained by transmission by asymptomatic people.

### The gold standard: rRT-PCR on nasopharyngeal swabs

The need for an accurate and rapid diagnosis has always represented a primary concern. At present, nasopharyngeal (NP) swabbing, followed by nucleic acid amplification tests (NAATs), in particular real time reverse transcription polymerase chain reaction (rRT-PCT), is recognised as the gold standard for the detection of SARS-CoV-2 infection (4). However, in many countries and settings, access to this type of testing has been increasingly difficult as it requires trained staff with personal protective equipment, health care resources and facilities, and is poorly accepted by patients as NP sampling tends to cause discomfort and sometimes bleeding. Furthermore, the risk for disease transmission to healthcare personnel when collecting samples is not negligible and the need for sample transportation may delay the results and, in turn, the time for diagnosis. In addition, over 60% of laboratories worldwide declared facing serious problems in obtaining reagents and kits for molecular testing (5). Putting together these limitations and considering the invasive nature of the procedure, self-collection of mid-nasal swabs have been proposed to increase testing access and minimize exposure risk to health care workers as well as the saving of personal protective equipment (6). However, self-collection of NP does not seem to overpass current bottlenecks for the likely low quality of sampling and some possible traumatic adverse events. In addition, self-collected samples require transport to a centralized laboratory, which often delay results for more days, drives down testing frequency and a timeless diagnosis.

### An alternative testing regimen

More recently, the evidence of a continuous increase of the number of infected patients and the fear for a further extent and spread of the pandemic which should constitute a “second wave of pandemic”, raised new efforts to change current scenario and to propose a new and improved strategy for containment. According to the recently published paper by Mina and Colleagues (7), the “key question is how effectively infections can be detected in a population by the repeated use of a given test as part of an overall strategy”. The Authors, therefore, invite the scientific community to “shift our attention from a narrow focus on the analytical sensitivity of a test to the more relevant measure of a testing regimen’s sensitivity to detect infections” (7). This should be done by adopting cheap and easy to perform testing, that allows repeated self-collection and analyses at home, using methods with a lower analytical sensitivity, even analytical limits 100 or 1000 times higher than rRT-PCR, but able to identify people “who are currently transmitting the virus” (7). According to this proposal, therefore, a regimen of regular testing should work as a filter through the identification, isolation and further tracing of contacts of all currently infected persons, including pre-symptomatic and asymptomatic subjects. The rationale of the new strategy is based on the evidence that current testing regimens have a sensitivity of 10% or even lower to detect infections and are failing to provide an efficient filter for COVID-1 (7). In addition, more than 50% of positive subjects identified by rRT-PCR-based surveillance had cycle threshold (CT) values higher than 30, indicating low viral RNA counts that have been associated with very low or absent cytotoxicity in vitro and no infective potential (8, 9).

Based on the previously reported rationale, Mina and colleagues are proposing the adoption of rapid lateral-flow antigen tests that are “cheap, produced in the tens of millions or more per week, and could be performed at home, opening the door to effective Covid filter regimens” (7). Recently, the Cochrane database reported data on the accuracy of currently available rapid antigen tests (COVID-19 Ag-RDTs) (10) and on September 11^th^, this year, WHO released an Interim guidance highlighting potential usefulness, limitations and appropriate scenarios for use COVID-19 Ag-RDTs stressing the need to adopt tests that meet the minimum performance requirements of ≥ 80% sensitivity and >97% specificity compared to a NAAT reference assay (11). The main question seems to be not the ability to detect Ct higher than 33 values, but “to perform well in patients with high viral load (Ct values < 25 or >10^6^ genomic virus copies/mL) which usually appears in the pre pre-symptomatic and early symptomatic phases of the disease (within the first 5-7 days of illness)” (7). However, Liotti and colleagues have found that the percent positivity agreement of a commercially available RDT with NAAT is >95% in upper respiratory tract specimens with high viral load (i.e. cycle threshold<25), but dramatically declines to 20-40% in samples with Ct >25 (12). Similar data have been reported by Porte and colleagues who also confirmed the evidence that diagnostic sensitivity is strongly dependent to the viral load (13). According to the systematic review already mentioned and prepared by the Cochrane COVID-19 Diagnostic Test Accuracy Group, the diagnostic accuracy of Ag-RDTs has been found to vary between 8-72% in samples with low viral load, being always lower than the WHO recommended value (>80%), thus stressing the need for a careful validation of the RDTs diagnostic performances to assure an appropriate usage of the innovative test regimen (10). In other words, the question if SARS-CoV-2 antigen assays could be safely and effectively used at home “like pregnancy tests” (14) cannot be answered, at least right now. Finally, if some reports have failed to demonstrate a detectable cytopathic effect in cultured cells in samples with Ct values higher than 33-35, this finding has been observed in a limited number of cases and requires more robust evidence collected on a large number of COVID-19 patients.

### Saliva as an alternative sample

However, our main concerns on the new testing regimen based on Ag-RDTs is that it still remains based on NP swabs. This represents a major limitation both for the invasive nature of this type of samples, particularly for children and patients with major neurodevelopmental and psychiatric issues, and the need for dedicated staff and settings, thus mitigating the analytical but not the logistical burden associated with a timely identification of cases. Evidence has been collected to demonstrate that saliva-based testing may be an alternative to the more widely used NP and oropharyngeal swabs for COVID-19 diagnosis and disease monitoring. In a recently published review, Sapkota and colleagues have reported the data of 5 already published papers which demonstrate the comparability of rRT-PCR results between NP swabs and salivary samples, that have been reported to be more consistent and sensitive than traditional swab samples (15). In particular, Wyllie and co-workers (16) have stressed the point that not only collection of saliva is more acceptable to patients as less invasive and that samples by patients themselves reduces the need for direct interaction with health care workers thus alleviating demands for supplies of personal protective equipment, but also that, in addition, the temporal profile of SARS-CoV-2 in saliva is more consistent as compared to NP swabs, thus allowing a more frequent testing regimen (7). Up to now, comparative studies between salivary and NP samples have been performed using rRT-PCR to detect SARS-CoV-2, but preliminary data collected using novel antigen-based tests are very promising, allowing the achievement of the analytical performances recommended by WHO (17). We recently evaluated the diagnostic performances of a quantitative antigen test for COVID-19 infection, which is a chemiluminescent assay (Lumipulse G SARS-CoV-2 Antigen, Fujirebio, Japan) automated on the Lumipulse G1200 analyzers, using salivary specimens. The method has been extensively and previously validated on NP swabs exhibiting a 99.6% specificity and 91.4% overall agreement rate (286/313) on the Lumipulse G600II automated immunoassay analyzer. In specimens with >100 viral copies and between 10 and 100 copies, the antigen test showed 100% and 85% concordance with rRT-PCR (18).

## Materials and Methods

A total of 40 patients (23 males, 17 females, age range 12-84 years), who were admitted to the Padova University hospital for COVID-19, were included in this study. For all individuals, a NP swab and a salivary sample were simultaneously collected 1 to 20 days after hospitalization. All NP samples were obtained using cotton swabs (ESwab Liquid Amies Medium). Patients were also asked to collect a salivary sample by using the Salivette device (Sarstedt, Germany), and invited to chew the cotton swab for at least one minute. Saliva was obtained after centrifugation at 3000 g for 5 minutes. Fresh NP swabs and salivary samples were analysed within 3 hrs from the collection. rRT-PCR was used to detect SARS-CoV-2 from NP swab using the diagnostic system TaqPath COVID-19 RT-PCR kit (Applied Biosystems, USA), which analyses ORF1 ab, N and S SARS-CoV-2 genes. Briefly, RNA was extracted using an automated platform (Magna Pure 96 Instrument, Roche Diagnostics, USA) and then used for rRT-PCR, which was performed by ABI prism® QuantStudio™ 5 Real-Time PCR Systems (Applied Biosystems, USA). RNaseP was analyses separately by QuantStudio™ 5 Real-Time PCR Systems (Applied Biosystems, USA) as previously described (19). SARS-CoV-2 Ag testing was performed on Lumipulse G1200 (Fujirebio, Japan) using 200 µL of saliva, mixed with 200 uL of Lumipulse G buffer.

## Results

Figure 1 shows the individual values of salivary SARS-CoV-2 antigen after subdividing patients on the basis of NP swab results obtained at the same time of saliva collection. The dotted line shows the cut-off suggested by the manufacturer (0.67 ng/L). Using this threshold, the sensitivity of salivary antigen was 62%, being the specificity 100%. By ROC curve analysis (mean ± SE area: 0.909 ± 0.05) the highest likelihood ratio fitted with the cut-off of 0.11 ng/L (continuous line in Figure 1). By using this revised threshold, sensitivity and specificity of salivary antigen were 71% and 97% respectively. Interestingly the 8 patients with very high levels of salivary antigen (above 3 ng/L) had a very short hospital stay (from 1 to 3 days, median 2 days), suggesting that salivary antigen testing might be highly effective for the rapid identification of new-onset SARS-CoV-2 infections and for screening purposes.

**Figure 1.**
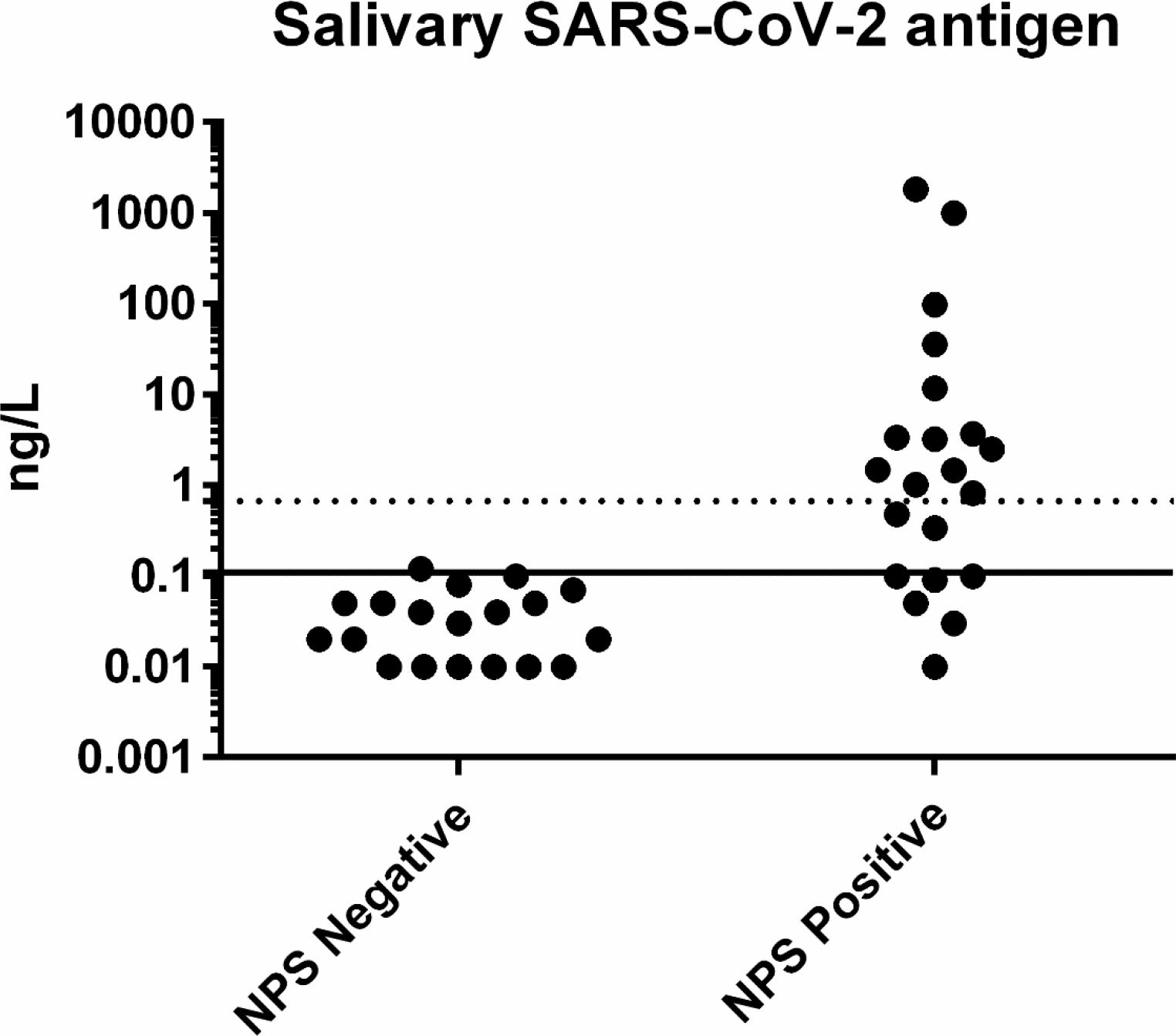
Individual values of SARS-CoV-2 antigen measured in saliva samples of COVID-19 inpatients. Patients are subdivided on the basis of negative or positive results of nasopharyngeal swabs (NPS) performed at the same time of saliva collection. Dotted line: manufacturer’s suggested cut-off (0.67 ng/L). Continuous line: ROC curve calculated cut-off (0.11 ng/L).

## Conclusions

As a series of flow-later immunochromatographic assays for rapid antigen detection have been already and will be developed, we would like to make a proposal that, adopting saliva as the sample of choice should be split into two different propositions according to the different setting and population to be tested.

The first proposition is the use of the automated quantitative antigen test using the Lumipulse G1200 which allows a rapid answer (30 seconds) and valuable throughput (120 tests/hour). This proposition should be performed to substitute/integrate molecular testing in centralized laboratories which may experience difficulties in reagent supply as well as in complying with valuable turnaround times. However, the most intriguing aspect is to use this proposition in airports, cruise ships and other settings in which there is the need to detect the infection and the risk of transmission of the virus with accuracy and rapidity, but do not allow further and frequent testing.

The second option is to adopt a diagnostic system which use salivary samples and easy to perform lateral-flow or similar devices which may allow a safe, self-testing to be performed at home, in closed or semi-closed groups such as schools, care-homes, work-places and dormitories. This option requires a well-defined test frequency which should allow the safely detection of infections and better fits with the strategy of containment proposed by Mina and Colleagues (7), but using salivary specimens instead of NP swabs, opening the door to an effective COVID-19 filter regimen. Of course in case of self-testing there are still some important issues to be solved such as tracing and reporting of results to a central public health authorities as well the implementation of effective preventive behaviour in case of positive results and further operational and social science studies are needed before widely recommending self-testing.

The third point is to adopt saliva testing as a candidate method for surveillance programs in subjects confined at home (for early interruption of quarantine). This proposal is supported by the easy and comfortable procedure and its suitability in outpatient settings.

A better knowledge of the true analytical and diagnostic performances (sensitivity first, and specificity too) of the testing devices as well as on the recommended testing frequency should be rapidly collected to allow the rationale introduction of the new containment strategy to ultimately measure its effectiveness and outcomes.

## Data Availability

Data is owned by the Authors. However, data could be shared if requested, despite the final decision of share results remain of Authors

